# Does earlier diagnosis and treatment of brain tumours matter? Time-to-treatment intervals and tumour size at detection; impact on survival, recurrence, inpatient length of stay and neurological deficit

**DOI:** 10.1101/2025.06.23.25330109

**Authors:** Ewan Gray, James Cameron, Abigail Lishman, Peter Hall, Piyumanga Karunaratne, Giovanni Tramonti, Maheva Vallet, Luke Pike, Matthew Baker, Paul M Brennan

## Abstract

**Background:** The importance of timely diagnosis of brain tumours is well recognised but how this translates to impacts on important patient outcomes is not well described. This study aimed to quantify the effect of time-to-treatment interval and tumour size at diagnosis on six patient outcomes: tumour-specific survival, overall survival, new or worsened neurological deficit, 30-day mortality, recurrence and inpatient length of stay in 12 months following diagnosis.

**Methods:** A retrospective cohort study including 1196 patients from Southeast Scotland diagnosed with brain tumours between 2010 and 2020. Regression methods (Cox, Logistic & Linear) used to estimate per day impact of time-to treatment and tumour size impact on outcomes.

**Results:** The mean time from first presentation to treatment was 161 days (median: 57 days) with wide variance observed (standard deviation: 330 days). Time-to-treatment results showed an association between longer intervals and reduced mortality. The mean tumour size was 4.06cm (SD: 1.75cm). Each 1cm increase in tumour size increased tumour-specific mortality risk by approximately 11% (HR: 1.11, 95%CI: 1.06-1.16), increased all-cause mortality by 6% (HR: 1.06, 1.02-1.11), increased the expected inpatient stay by 1.5 (0.2-2.8) days, and the odds of new or worsened neurological deficit by 11% (OR: 1.11, 1.01-1.21).

**Conclusions:** Larger tumour size was consistently associated with increased hazard ratios for mortality. Applying these estimates together with estimates of mean tumour growth rates available in the literature, if a 6cm Glioblastoma tumour were diagnosed 1 month earlier then we would expect a 18-28% reduction in hazard of brain tumour mortality.

**Funding:** Dxcover Ltd.

**Research in Context:** *Evidence before this study:* Brain tumour is frequently diagnosed in an emergency setting, with multiple primary care visits often preceding a diagnosis, suggesting opportunities for earlier diagnosis. Previous studies have produced conflicting findings regarding the impact of diagnostic delays on survival and other patient outcomes. Some suggest that longer intervals are associated with worse prognosis, while others report a paradoxical association where delayed diagnosis appears to correlate with improved survival, the ‘waiting time paradox’. Prior research has largely focused on the prognostic value of tumour size rather than its role as a mediator of diagnostic delay, limiting causal inference regarding the benefits of earlier detection.

*Added value of this study:* This study addresses gaps in the literature by examining tumour size as a key mediating factor in the relationship between diagnostic timing and patient outcomes. Using a large, well-curated dataset of 1,196 patients from Southeast Scotland, we demonstrate that tumour size at diagnosis is strongly associated with mortality risk, neurological deficits, inpatient stay, and recurrence. A one-month earlier diagnosis of a 6cm glioblastoma, for example, could reduce tumour size sufficiently to lower the hazard of brain tumour mortality by 18– 28%. By disentangling tumour size effects from time-to-treatment intervals, our study provides a clearer framework for evaluating early detection strategies in neuro-oncology.

*Implications of all the available evidence:* These findings reinforce the potential impact of earlier diagnosis for brain tumour patients. Building upon the accumulated evidence that tumour size is a critical factor influencing prognosis, this study is the first to robustly estimate the causal association. Earlier detection, through influence on tumour size at diagnosis, may offer measurable survival benefits and potential reductions in healthcare resource use. Given the observational nature of this study, further prospective trials and mathematical modeling are needed to assess the impact of early diagnosis interventions at a population level.

## Introduction

Diagnosis of brain tumours suffers from two related problems, an excess of emergency presentations (1) and long diagnostic intervals. (2) NHS England route-to-diagnosis data show that approximately 40% of primary brain cancers present as an emergency each year (1) In the UK, most (80%) brain tumour patients report one or more primary care visit in the months preceding diagnosis (3), and more than a third of patients have 3 or more primary care visits recorded (4). These data suggest opportunities for earlier diagnosis (5).

Despite the potential for earlier diagnosis, interventions aiming to improve speed and accuracy of diagnosis face barriers in demonstrating their impact and value. While the importance of a timely diagnosis is recognised by professionals and policy makers, how this translates in a quantitative sense to important patient outcomes such as survival, quality of life and length of hospital stays, is not well described. Consequently, the impact of earlier diagnoses on population health and healthcare resource use are highly uncertain. This creates difficulty for the economic evaluation of interventions aiming to aid earlier brain tumour diagnosis (6,7). Evidence from real world observational data can help address this.

The Edinburgh brain tumour database utilised by this study is well suited to interrogate the relationship between time to diagnosis and patient outcomes. This manually curated data was collected specifically to understand clinical pathways for brain tumour patients and can be supplemented through linkage to routine healthcare records.

Prior studies in this area have explored only limited aspects of the relationship between earlier (or delayed) diagnosis, treatment, and patient outcomes. The overall pattern of findings reveals inconsistency, with positive (longer times associated with improved outcome) (8), negative (9,10) and null (11) associations identified. An important part of the explanation of these seemingly contradictory findings is the ‘waiting time paradox’ (12–14), the phenomenon where a delayed diagnosis or treatment can paradoxically mean a better prognosis for the patient. This is due to differences between cases in clinical features that may induce delay and are also associated with better outcomes. For example, a longer diagnostic process is more likely for less severely impairing symptoms, and such symptoms may be more commonly associated with less aggressive disease.

To better elucidate the relationship between earlier or delayed diagnosis and patient outcomes, this study investigated two associations. Firstly, the association between time-to-treatment and outcomes. Secondly, the association between tumour size and outcomes.

Tumour size is expected to mediate the impact of earlier or delayed diagnosis through the process of tumour growth. Pre-operative tumour size is a well-defined prognostic factor for both low- and high-grade brain tumours (15–19). Larger tumour size can increase the risk of both neurological deficits at presentation and surgical complications following resection.

While the relationship between tumour size and patient outcomes is well described in the literature, this study examines it from a unique perspective. Prior studies were concerned with the prognostic (predictive) value of tumour size rather than trying to estimate a causal effect (20). The multivariate analysis reported in these studies are inappropriate from a causal perspective due to including potential confounders such as tumour growth rate (21), mediators of the effect of tumour size such as extent of resection(16,17), or potential outcomes of tumour size (colliders) such as pre-operative symptoms (17,18). The reported results are therefore potentially biased estimates of the causal effect of tumour size on patient outcomes.

Patient outcome variables were selected based on known clinical relevance and availability in the database. The main objectives of the study were specified as estimation of the association between the two indicators of earlier or later diagnosis, i) tumour size and ii) length of the presentation to treatment interval, with the patient outcomes of brain tumour specific survival and overall survival. Additional objectives were to estimate the association of these indicators with the patient outcomes of new or worsened neurological deficit, 30-day mortality, cancer recurrence and inpatient length of stay in 12 months following diagnosis.

## Methods

### Study design

A retrospective cohort study.

### Patient data

All patients were identified from the Edinburgh brain tumour database, which includes consecutive patients whose care was discussed at the institutional neuro-oncology multi-disciplinary meeting. Patients diagnosed with brain tumours between January 1^st^ 2010 to 31 May 2020 were included in the study. Follow up was available until 31^st^ of May 2021 for all patients.

Brain tumour diagnosis was defined as a radiological diagnosis with or without recorded pathology diagnosis. Brain tumour subtypes were defined based on diagnosis labels manually recorded form the radiology report. Inclusion criteria limited the study to patients aged 18 and over. Exclusion criteria of previous history of other cancer within the last 3 years was applied.

### Data processing

Patient information was collected from manually curated fields compiled in the database and supplemented by data-linkage with national routine datasets (SMR00 - Outpatient, SMR01-Inpatient and SMR06 – Cancer Registry). Pseudonymous data linkage was achieved by linking the patient’s database id number to their NHS Scotland Community Health Index (CHI) number. Data processing was completed within an NHS IT environment by an NHS analyst.

Tumour size was defined as the largest diameter of the tumour given in the diagnostic report (CT or MRI). Time-to-treatment interval was defined in two ways, [1] as time from first presentation to first recorded treatment and [2] time from radiological diagnosis to first recorded treatment. Time of first presentation was defined as the date of first recorded clinical appearance or the first date of recorded symptoms if this was earlier. Date of diagnosis refers to the date of the first imaging procedure that confirmed a radiological diagnosis. The date of treatment is the date first treatment was received, whether this was surgery, radiotherapy or chemotherapy.

Outcome variables included all-cause mortality, brain tumour specific mortality (Supplementary Appendix reports ICD-10 codes included for this outcome), 30-day (all-cause) mortality, new or worsened neurological deficit, tumour recurrence, and total inpatient bed days within 12 months of the date of diagnosis. Brain tumour specific mortality and all-cause mortality were selected as the main outcomes of interest. While other outcomes were considered additional items to supplement these most clinically relevant outcomes.

Other demographic and clinical factors available for the descriptive analysis included age at diagnosis, sex, tumour location, tumour grade, route of presentation, Charlson comorbidity index, deprivation level (SIMD quintile), urban/rural residence. Treatments received were recorded for the categories of surgery, radiotherapy, and use of temozolomide.

### Statistical analysis

Mortality outcomes analysed by survival analysis were considered the primary outcomes. Hazard ratios were estimated for cancer-specific, overall mortality and tumour recurrence using Cox regression, including covariates to adjust estimates for potential confounding variables. For the additional outcomes of new or worsened neurological deficit, and 30-day mortality, odds ratios were estimated by logistic regression. Linear regression was used to estimate effects on the number of inpatient bed days. This was subject to first identifying if a linear relationship offered a reasonable approximation based on examining scatterplots and QQ plots.

The covariates included in all Cox, Logistic, or Linear regressions were age at diagnosis, sex, deprivation level (SIMD quintile), Charlson comorbidity index, and Glioblastoma (binary indicator, only applies for the pooled cohort).

After examining the time-to-treatment data distributions and associations it was noted that interpretation was made challenging due to the presence of outliers. This caused estimated coefficients to be extremely small and in many cases indistinguishable from zero up to the fourth decimal place. To address this, a post-hoc analysis which stratified on time-to-treatment, using clinically relevant categories was added. The categories applied were <2 weeks, 2-6 weeks, 6-12 weeks and >12 weeks.

Complete case analysis was conducted therefore patients with missing data in relation to predictor, covariate or outcomes variables were excluded from the regression analysis.

### Tumour Sub-type Stratified Analysis

Analysis was repeated identically, limited to patients in each of three principal brain tumour subtype categories. This was conducted with the intention explore the consistency of effects across subtypes. Tumours were grouped as Glioblastoma (GBM), Non-glioblastoma primary brain tumours (Non-GBM PBC), and Brain metastases. The categories were chosen based on clinical relevance while still being sufficiently broad to allow meaningful results. Note that the metastases group included in the database was intentionally limited to exclude those with known active primary tumours at other sites (i.e. it includes those that presented with a brain tumour subsequently discovered to be secondary to another site).

### Role of the funding source

Dxcover Ltd provided funding for data access and analysis. The study design was created independently and subsequently approved by the funder.

## Results

1196 of 1437 patients entered in the database were included in the study (Figure 1), comprising of 459 patients with Glioblastoma, 587 with other primary brain tumours and 150 with brain metastases. Descriptive statistics are displayed in Table 1. The mean age was 63. The mean tumour size was 40.6mm (SD: 17.5mm). Tumours were most commonly located in the frontal lobe (36.3%). Tumour size data was missing for 126 patients.

**Fig. 1.**
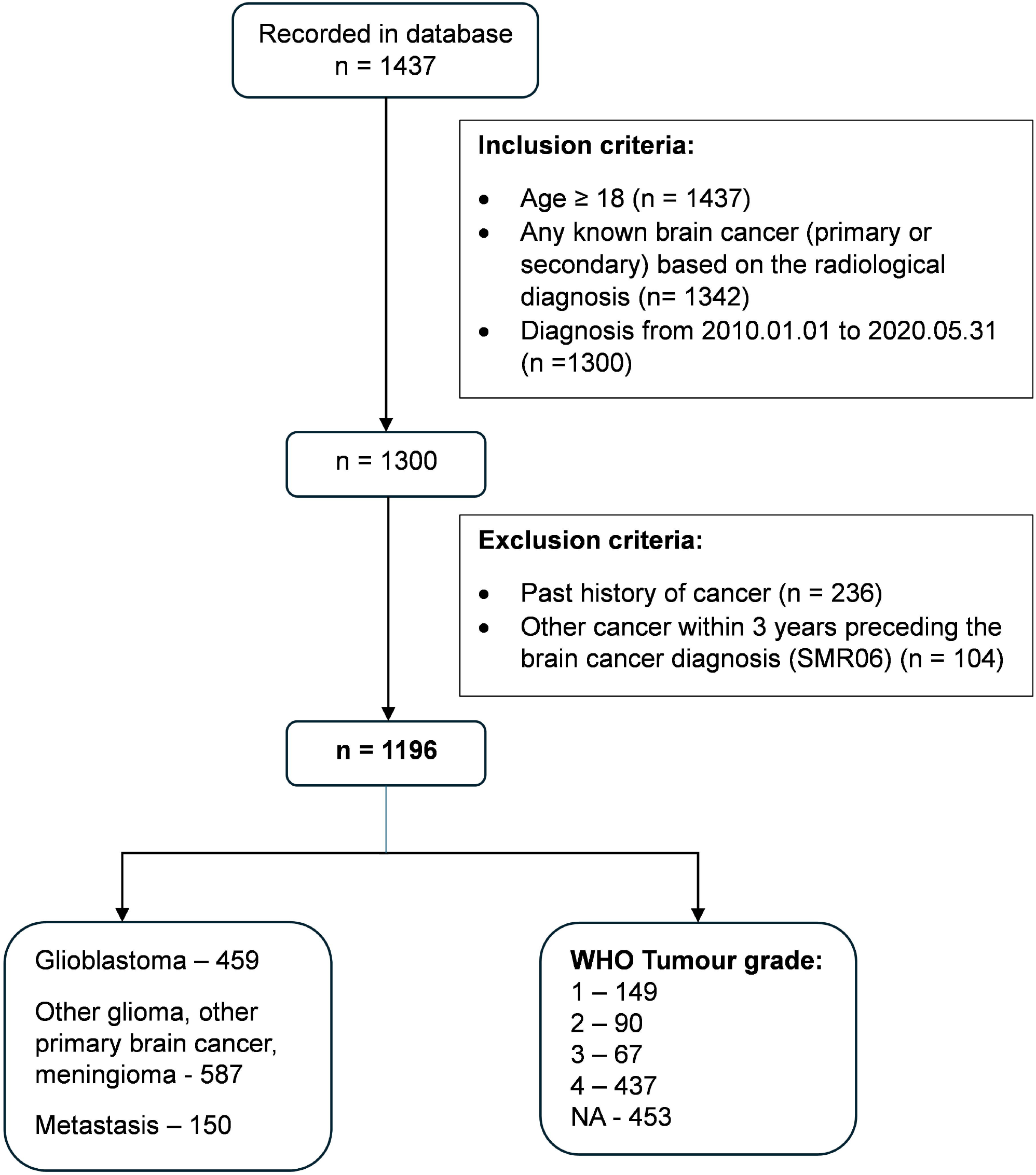

**Table 1.** Descriptive Statistics.

|  | **Glioblastoma**<br>**(n = 459)** | **Non-GBM**<br>**Primary**<br>**Tumour (n =**<br>**587)** | **Brain**<br>**metastases**<br>**(n = 150)** | **Overall**<br>**(N=1196)** |
| --- | --- | --- | --- | --- |
| **Age at diagnosis** |  |  |  |  |
| Mean (SD) | 62.9 (13.1) | 55.5 (16.6) | 64.0 (11.4) | 59.4 (15.2) |
| Median (Q1, Q3) | 65.0 (54.0, 72.0) | 57.0 (43.0, 68.0) | 65.0 (56.2, 72.0) | 61.0 (50.0, 70.0) |
| **Sex** |  |  |  |  |
| Female | 185 (40.3%) | 316 (53.8%) | 75 (50.0%) | 576 (48.2%) |
| Male | 274 (59.7%) | 271 (46.2%) | 75 (50.0%) | 620 (51.8%) |
| **Primary cancer subtype** |  |  |  |  |
| Glioblastoma | 459 (100.0%) | 0 (0.0%) | 0 (0.0%) | 459 (38.4%) |
| Other Glioma | 0 (0.0%) | 293 (49.9%) | 0 (0.0%) | 293 (24.5%) |
| Meningioma | 0 (0.0%) | 178 (30.3%) | 0 (0.0%) | 178 (14.9%) |
| Other primary brain cancer | 0 (0.0%) | 116 (19.8%) | 0 (0.0%) | 116 (9.7%) |
| Metastasis | 0 (0.0%) | 0 (0.0%) | 150 (100.0%) | 150 (12.5%) |
| **Tumour size at diagnosis (cm)** |  |  |  |  |
| Mean (SD) | 4.7 (1.6) | 3.8 (1.8) | 3.0 (1.2) | 4.1 (1.8) |
| Median (Q1, Q3) | 4.7 (3.5, 5.9) | 3.6 (2.4, 5.0) | 3.0 (2.2, 3.7) | 4.0 (2.8, 5.3) |
| Number Missing | ≥30 | 87 | <10 | 126 |
| **Tumour size category (cm)** |  |  |  |  |
| < 3 | 51 (12.0%) | 173 (34.6%) | 69 (47.9%) | 293 (27.4%) |
| 3 – 5.9 | 271 (63.6%) | 257 (51.4%) | 73 (50.7%) | 601 (56.2%) |
| ≥ 6 | 104 (24.4%) | >50 (>5%) | <10 (<5%) | 176 (16.4%) |
| Number Missing | ≥30 | >50 | <10 | 126 |
| **Tumour location** |  |  |  |  |
| Frontal | 166 (37.4%) | 191 (36.9%) | 46 (30.9%) | 403 (36.3%) |
| Temporal | 104 (23.4%) | 95 (18.3%) | 11 (7.4%) | 210 (18.9%) |
| Parietal | 72 (16.2%) | 72 (13.9%) | 20 (13.4%) | 164 (14.8%) |
| Occipital | ≥10 (≥2.3%) | ≥10 (≥1.9%) | <10 (<6.7%) | 36 (3.2%) |
| Cerebellar | <10 (<2.3%) | ≥10 (≥1.9%) | 33 (22.1%) | 64 (5.8%) |
| Brainstem | <10 (<2.3%) | 33 (6.4%) | <10 (<6.7%) | 37 (3.3%) |
| Multiple | 86 (19.4%) | 79 (15.3%) | 32 (21.5%) | 197 (17.7%) |
| Number Missing | ≥10 | 69 | <10 | 85 |
| **Tumour grade** |  |  |  |  |
| 1 | <10 (<3.1%) | 143 (37.1%) | <10 (<33.3%) | 149 (20.1%) |
| 2 | <10 (<3.1%) | 87 (22.6%) | <10 (<33.3%) | 90 (12.1%) |
| 3 | <10 (<3.1%) | 62 (16.1%) | <10 (<33.3%) | 67 (9.0%) |
| 4 | 320 (97.6%) | 93 (24.2%) | 24 (80.0%) | 437 (58.8%) |
| Number Missing | 131 | 202 | 120 | 453 |
| **Presentation** |  |  |  |  |
| Primary care | 20 (6.6%) | 42 (9.5%) | 14 (11.1%) | 76 (8.7%) |
| Secondary care | 161 (53.5%) | 245 (55.2%) | 61 (48.4%) | 467 (53.6%) |
| Emergency | 120 (39.9%) | 157 (35.4%) | 51 (40.5%) | 328 (37.7%) |
| Number Missing | 158 | 143 | 24 | 325 |
| **Treatment received**<br>**(Surgery, Radiotherapy and/or Temozolomide)** |  |  |  |  |
| Yes | 355 (77.3%) | 453 (77.2%) | 104 (69.3%) | 912 (76.3%) |
| No | 104 (22.7%) | 134 (22.8%) | 46 (30.7%) | 284 (23.7%) |
| **Surgery** |  |  |  |  |
| Yes | 334 (72.8%) | 437 (74.4%) | 87 (58.0%) | 858 (71.7%) |
| No | 125 (27.2%) | 150 (25.6%) | 63 (42.0%) | 338 (28.3%) |
| **Radiotherapy** |  |  |  |  |
| Yes | 309 (67.3%) | 269 (45.8%) | 67 (44.7%) | 645 (53.9%) |
| No | 150 (32.7%) | 318 (54.2%) | 83 (55.3%) | 551 (46.1%) |
| **Temozolomide** |  |  |  |  |
| Yes | 199 (43.4%) | 114 (19.4%) | 17 (11.3%) | 330 (27.6%) |
| No | 260 (56.6%) | 473 (80.6%) | 133 (88.7%) | 866 (72.4%) |
| **Comorbidity (CCI)\*** |  |  |  |  |
| 0 | 391 (85.2%) | 522 (88.9%) | 96 (64.0%) | 1009 (84.4%) |
| 1-2 | 52 (11.3%) | 53 (9.0%) | 18 (12.0%) | 123 (10.3%) |
| 3 or more | 16 (3.5%) | 12 (2.0%) | 36 (24.0%) | 64 (5.4%) |
| **Deprivation score (SIMD)\*\*** |  |  |  |  |
| 1 (Most deprived) | 59 (12.9%) | 68 (11.7%) | 18 (12.1%) | 145 (12.2%) |
| 2 | 103 (22.5%) | 132 (22.7%) | 29 (19.5%) | 264 (22.2%) |
| 3 | 91 (19.9%) | 111 (19.1%) | 39 (26.2%) | 241 (20.3%) |
| 4 | 103 (22.5%) | 134 (23.1%) | 28 (18.8%) | 265 (22.3%) |
| 5 (Least deprived) | 102 (22.3%) | 136 (23.4%) | 35 (23.5%) | 273 (23.0%) |
| Number Missing | <10 | <10 | <10 | <10 |
| **Urban/Rural residence** |  |  |  |  |
| Urban | 327 (76.0%) | 419 (81.7%) | 108 (80.0%) | 854 (79.2%) |
| Rural | 103 (24.0%) | 94 (18.3%) | 27 (20.0%) | 224 (20.8%) |
| Number Missing | 29 | 74 | 15 | 118 |
| **Healthcare Resource Use** |  |  |  |  |
| **Outpatient visits** |  |  |  |  |
| Mean (SD) | 4.3 (3.1) | 5.3 (3.7) | 6.1 (5.0) | 5.0 (3.7) |
| Median (Q1, Q3) | 4.0 (2.0, 6.0) | 5.0 (3.0, 7.0) | 5.0 (2.0, 9.0) | 4.0 (2.0, 7.0) |
| Number Missing | 74 | 58 | 23 | 155 |
| **Inpatient visits** |  |  |  |  |
| Mean (SD) | 5.2 (3.0) | 4.9 (3.1) | 6.0 (3.8) | 5.2 (3.2) |
| Median (Q1, Q3) | 5.0 (3.0, 7.0) | 4.0 (3.0, 6.0) | 5.0 (3.0, 7.0) | 4.0 (3.0, 7.0) |
| Number Missing | 31 | 103 | 11 | 145 |
| **Total length of stay (days)** |  |  |  |  |
| Mean (SD) | 32.1 (33.1) | 28.6 (38.0) | 34.5 (32.4) | 30.8 (35.4) |
| Median (Q1, Q3) | 21.0 (8.0, 47.0) | 13.0 (6.0, 38.0) | 24.0 (13.0, 48.0) | 18.0 (7.0, 44.0) |
| Number Missing | 31 | 103 | 11 | 145 |
| **Average length of stay (days)** |  |  |  |  |
| Mean (SD) | 6.6 (6.5) | 5.6 (6.3) | 6.7 (7.2) | 6.1 (6.5) |
| Median (Q1, Q3) | 4.4 (2.0, 9.0) | 3.0 (1.8, 7.3) | 4.7 (2.4, 7.5) | 3.8 (2.0, 7.9) |
| Number Missing | 31 | 103 | 11 | 145 |
| **Time radiological diagnosis to first treatment (days)** |  |  |  |  |
| Mean (SD) | 23.4 (47.4) | 149.8 (361.5) | 41.5 (123.8) | 88.3 (267.0) |
| Median (Q1, Q3) | 16.0 (9.5, 25.5) | 37.0 (14.0, 114.0) | 19.0 (12.0, 36.0) | 21.0 (12.0, 49.0) |
| Number Missing | 104 | 134 | 47 | 285 |
| **Time from first presentation to treatment (days)** |  |  |  |  |
| Mean (SD) | 62.6 (79.6) | 260.0 (438.1) | 73.1 (132.6) | 161.3 (329.7) |
| Median (Q1, Q3) | 39.0 (24.5, 70.0) | 99.0 (40.0, 266.0) | 40.0 (27.0, 70.0) | 56.5 (30.0, 132.2) |
| Number Missing | 116 | 156 | 52 | 324 |
| **Time from primary care referral to treatment (days)** |  |  |  |  |
| Mean (SD) | 31.9 (58.1) | 177.5 (335.1) | 35.4 (44.6) | 105.1 (250.4) |
| Median (Q1, Q3) | 18.0 (11.0, 33.2) | 56.0 (19.0, 177.0) | 22.0 (14.0, 41.2) | 28.0 (13.5, 82.5) |
| Number Missing | 219 | 268 | 70 | 557 |
| *Note: Small numbers are censored due to information governance requirements of the source data.* |  |  |  |  |

The most common route to diagnosis was via the emergency department (37.7%) and the majority of patients received surgical treatment (72%). In addition, 54% received radiotherapy treatment and 28% recorded use of temozolomide.

Survival varied by tumour subtype in the expected pattern. Median survival was 8.3 months for the GBM group, 94 months for non-GBM primary brain tumours and 7.6 months for metastases (Figure 2).

**Fig. 2.**
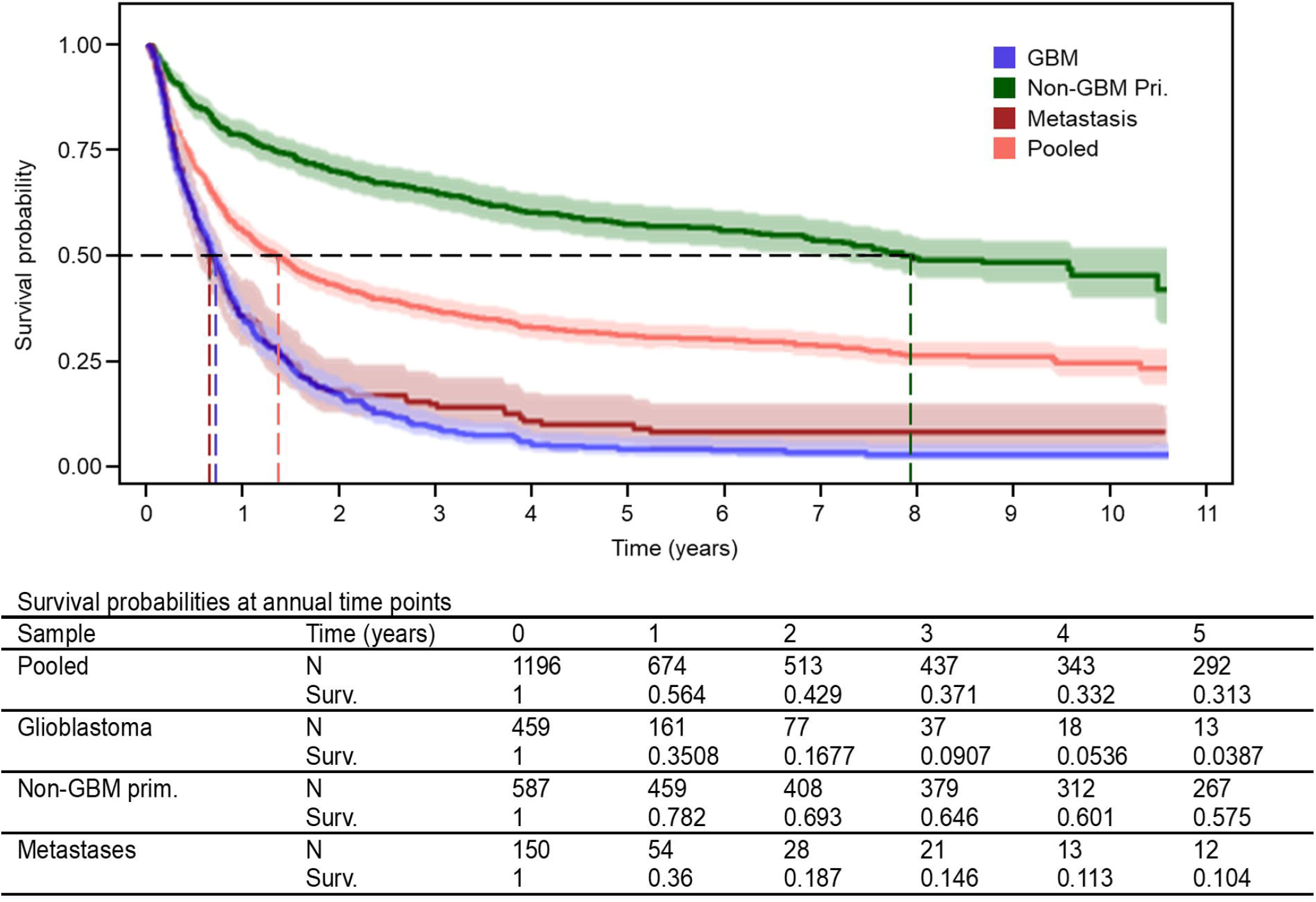

The time in clinical pathways patients followed from first presentation through referrals, diagnosis, and initial treatment are summarized in Table 1. The mean time from first presentation to treatment in the pooled sample was 161 days (median: 57 days) and was missing in 324 patients. Time interval varied by cancer subtype, for GBM patients the mean was 63 days (median: 39 days), for non-GBM PBC patients it was 260 days (median: 99 days) and for patients with metastases 73 days (median: 40 days). Means being higher than medians, and large standard deviations (80-438 days), indicate strongly right skewed distributions in all categories. Analysis of the difference in means between presentation to treatment and diagnosis to treatment indicates an average of around 40 days from presentation to diagnosis, with wide variation. High variance in these intervals reflects both variation in the duration until symptoms are recognised as a possible tumour, and variation in triage following diagnosis. For patients some with non-GBM primary brain tumours the variance is likely to also reflect watchful waiting strategies following diagnosis.

Associations between tumour size and patient outcomes are reported in Table 2. Tumour size (per cm larger) was associated with an increased risk of mortality from brain tumour (HR: 1.11, 95%CI: 1.06-1.16) and all-cause mortality (HR: 1.06, 95%CI: 1.02-1.11). These associations were observed to be consistent across subtypes (Figure SA1 & Table 2). The hazard ratio for tumour-specific mortality was 1.12 (95%CI:1.05 – 1.18) for GBM tumours, 1.13 (95%CI:1.04 – 1.22) for non-GBM primary tumours and 1.15 (95%CI:0.96 – 1.39) for metastases. Equivalent hazard ratios for all-cause mortality were 1.11 (1.05 – 1.17) for GBM tumours, 1.09 (1.02 – 1.17) for non-GBM primary tumours and 1.21 (1.02 – 1.42) for metastases.

**Table 2.** Tumour Size & Patient Outcomes Association (multivariable adjusted) *Reduced number of observations due to missing outcome data, number of observations given in square brackets.

| **Main Outcomes** | **Pooled cohort**<br>**(n = 1064)** | **GBM**<br>**(n = 397)** | **Non-GBM**<br>**Primary**<br>**Tumour**<br>**(n = 415)** | **Brain**<br>**metastases**<br>**(n = 132)** |
| --- | --- | --- | --- | --- |
| *Hazard Ratio, Odds Ratio, Coefficient (95% Confidence Interval)* |  |  |  |  |
| **Brain cancer mortality** | 1.11<br>(1.06 – 1.16) | 1.12<br>(1.05 – 1.18) | 1.13<br>(1.04 – 1.22) | 1.15<br>(0.96 – 1.39) |
| **All-cause mortality** | 1.06<br>(1.02 – 1.10) | 1.11<br>(1.05 – 1.17) | 1.09<br>(1.02 – 1.17) | 1.21<br>(1.02 – 1.42) |
| **Additional Outcomes** |  |  |  |  |
| **30-day mortality** | 1.17<br>(0.94 – 1.46) | 1.47<br>(1.06 – 2.05) | 0.89<br>(0.55 – 1.35) | 1.46<br>(0.83 – 2.70) |
| **Recurrence** | 1.00<br>(0.94 – 1.06) | 0.91<br>(0.83 – 1.00) | 1.15<br>(1.06 – 1.26) | 1.05<br>(0.81 – 1.35) |
| **New or worsened neurological deficit** | 1.11<br>(1.01 – 1.21) | 1.00<br>(0.87 – 1.15) | 1.24<br>(1.09 – 1.42) | 0.89<br>(0.56 – 1.38) |
| **Inpatient bed days, 12 months following diagnosis\*** | 1.50<br>(0.19 - 2.81)<br>[Obs. = 944] | 1.51<br>(-0.57 - 3.59)<br>[Obs. = 397] | 1.50<br>(-0.4 - 3.4)<br>[Obs. = 415] | 1.09<br>(-3.63 - 5.81)<br>[Obs. = 132] |

An association with increased risk of 30-day mortality was observed but this was not statistically significant in the pooled group (OR: 1.17, 95%CI: 0.94-1.46). In the GBM group the effect on 30-day mortality was statistically significant (OR: 1.47 95%CI: 1.06 - 2.05).

In the pooled group, 445 recurrence events were observed (37% of the sample). No association of size with risk of recurrence (OR: 1.00, 95%CI: 0.94-1.06) was observed. This may reflect opposing effects in the GBM group (HR: 0.91, 95%CI: 0.83-1.00) and non-GBM primary tumours (HR: 1.15, 95%CI:1.06-1.26).

New or worsened neurological deficit (OR: 1.11, 95%CI: 1.01-1.21) and longer total duration of inpatient stays (Coef. 1.50, 95%CI: 0.19 - 2.81) were also associated with larger tumour sizes. Compared to tumour-specific and all-cause mortality more variation and wider confidence intervals were observed across tumour-subtypes in relation to these outcomes (Table 2). Equivalent unadjusted (univariate) results for all associations of tumour size with outcomes, both in the pooled sample and stratified by tumour sub-type, are available in the Supplementary Appendix (Tables SA 1-4). Unadjusted results are very similar to the presented regression adjusted estimates with some small attenuation of effects following adjustment observed.

Increased time to treatment (per day) was associated with improved outcomes (Table 3) in relation to brain tumour mortality (HR: 0.9991, 95%CI:0.9985 - 0.9996), all-cause mortality (HR: 0.9985, 95%CI:0.998 - 0.9991), 30-day mortality (OR: 0.881, 95%CI: 0.689 - 0.975) and tumour recurrence (HR:0.9995, 95%CI:0.9991 - 0.9999). Note that the combination of using hazard ratios for a per day change, and existence of long intervals, cloud interpretation of clinical importance. To illustrate the potential clinical relevance, the per day hazard ratio of brain tumour mortality of 0.9991 in the pooled cohort translates to a hazard ratio of 0.85 for a 180-day longer interval.

**Table 3.** Time-to-treatment & Patient Outcomes Association.

| Main Outcomes | Pooled cohort<br>(n = 872) | GBM<br>(n = 342) | Non-GBM<br>Primary Tumour<br>(n = 428) | Brain metastases<br>(n = 97) |
| --- | --- | --- | --- | --- |
| *Hazard Ratio, Odds Ratio, Coefficient (95% Confidence Interval)* |  |  |  |  |
| **Brain cancer mortality** | 0.9991<br>(0.9985 - 0.9996) | 1.0001<br>(0.9985 - 1.0016) | 0.9991<br>(0.9985 - 0.9997) | 0.9974<br>(0.9921 - 1.0027) |
| **All-cause mortality** | 0.9985<br>(0.998 - 0.9991) | 1.0012<br>(0.9998 - 1.0027) | 0.9985<br>(0.9979 - 0.9992) | 0.9993<br>(0.9975 - 1.0012) |
| **Additional Outcomes** |  |  |  |  |
| **30-day mortality** | 0.881<br>(0.689 - 0.975) | 0<br>(0 - 5.043 x 10<sup>34</sup>) | 1<br>(0 - 6.489 x 10<sup>33</sup>) | 0.877<br>(0.4748 - 1.0386) |
| **Recurrence** | 0.9995<br>(0.9991 - 0.9999) | 0.9986<br>(0.9965 - 1.0007) | 0.9996<br>(0.9992 - 1.0000) | 0.9978<br>(0.9933 - 1.0023) |
| **New or worsened neurological deficit** | 1.0001<br>(0.9996 - 1.0006) | 0.9976<br>(0.9936 - 1.0007) | 1.0001<br>(0.9996 - 1.0006) | 0.9993<br>(0.9921 - 1.0031) |
| **Inpatient bed days, 12 months following diagnosis** | n/a | n/a | n/a | n/a |

No association with new or worsened neurological deficit or inpatient bed days was observed. The Supplementary Appendix provides results for unadjusted regressions (Tables SA 5,7,9,11), results for the alternative time-to-treatment definition (Tables SA 5-12), and results for the same analyses stratified by categories of time-to-treatment window (Tables SA 13-20).

Analysis stratified by time-to-treatment window categories (<2 weeks, 2-6 weeks, 6-12 weeks and >12 weeks) shows similar results to the base case analysis, improved outcomes with longer time windows, but with larger effect sizes observed in the 2-6 and 6-12 week groups (Tables SA 13-20).

Linear regression to estimate the relationship between time-to-treatment and inpatient length of stay was not applied because visual inspection of scatterplot and QQ plot did not suggest any approximately linear relationship was present.

## Discussion

Causal effects of earlier or later diagnosis of brain tumours are difficult to identify because experimental methods cannot be used, and observational data are challenging to collect and interpret. To overcome this difficulty, we have investigated the relationship between tumour size and patient outcomes as it is believed to play an important role mediating the effect of earlier or delayed diagnosis. This is predicated on a simple chain of causation. Tumours grow over time, and for brain tumours, size of the primary tumour has a very direct relationship with the severity of disease as well as the likelihood of successful treatment. Larger tumours will affect healthy brain tissues to a greater degree, compromising function, while complete and partial resections will carry greater risks of surgical complications. Delays to diagnosis allow greater time for tumour growth.

We have found evidence of the previously described ‘waiting time paradox’ in the relationship between time-to-treatment and patient outcomes. In common with previous investigators, we believe this is due to differences between cases in observed and unobserved prognostic factors. Prognostic factors affect how clinicians and patients assess the urgency of investigation and treatment. For example, in patients with similar symptoms, those whose symptoms progress more rapidly are likely to have diagnostic imaging earlier. Tumour growth and symptom progression is more likely with more aggressive tumours, which have a worse prognosis than lower grade tumours. Lower grade tumours grow more slowly, so symptoms will progress at a lower rate and diagnosis may therefore take longer, but these tumours have a better outcome. Hence, why for some patients a longer time to diagnosis is associated with a better outcome.

We have observed several features that should increase our confidence in a causal interpretation of the impacts of tumour size on the patient outcomes of survival, brain tumour specific or all-cause, new or worsened neurological deficit, and inpatient length of stay. These include a strong association, clear temporality, a gradient of effect, consistency and clinical plausibility (22). Firstly, a strong association was observed in relation to cancer-specific mortality, all-cause mortality, neurological deficit and length of inpatient stay. Temporality is clear as tumour size is recorded at the time of radiological diagnosis. The expected gradient was observed in relation to length of inpatient stay. The observed associations were consistent across subtypes in relation to cancer-specific and all-cause survival and were consistent for inpatient stays except for brain metastases. Finally, because the mainstay of treatment is surgical resection and, ceteris paribus, surgery becomes more difficult (and urgent) with increased tumour size, the case for clinical plausibility is strong.

To illustrate the potential importance of the relationship between time-to-diagnosis, tumour size and outcomes consider as an example a patient with a GBM 60mm diameter at diagnosis (75th percentile in this patient population). Applying two alternative mean growth rate estimates derived by previous studies (23,24), this tumour would be expected to be 31.0-45.5mm if diagnosed 1 month earlier. The reduction in tumour size would imply a 18-28% decrease in the hazard of brain cancer mortality. In this example that would translate to approximately 37-42% rather than 30% of patients surviving at 1 year, and 5-8% rather than 3% of patients surviving at 3 years following diagnosis. Inpatient bed days would be reduced from approximately 35.1 to 30.6-32.5 days, saving approximately £1300-2200 in healthcare expenditure(25) (details of calculations for this worked example are provided in the Supplementary Appendix).

The impact of such earlier detection for the whole population of brain tumour patients, across sub-types and size at diagnosis, could be predicted in a future modelling study, providing input for additional analysis of the overall impact of early diagnosis. This would support economic evaluation of interventions targeting earlier diagnosis.

An interesting further conclusion arising from this model of tumour growth and early detection is that because tumours grow approximately exponentially, the period of the last doubling in size prior to detection via symptoms is vitally important. This is when each day or week of earlier detection will have the largest effect on the tumour size at diagnosis. It follows that a few weeks can make a difference in the symptomatic setting while by the same logic it may also be true that a few weeks would be inconsequential in the setting of asymptomatic screening.

It is important to note the limitations imposed by the observational design of this study. Without having recourse to experimental data on delays to diagnosis and treatment, which could only become available through a natural experiment, uncertainty will remain about whether the observed associations are causal. More definitive evidence may require interventional studies aimed at early detection with new technologies by either modification of diagnostic pathways or by screening. These studies would be necessarily large, expensive and lengthy. Another limitation of this study is possible error in assigning the correct time of diagnosis or first presentation based on review of health care records. The true first presentation may not have been recognised or recorded within primary care. While we believe this does not have a major impact in these data if it were the case then this would have the effect of artificially shortening the observed intervals. Around ten percent of cases were excluded in the tumour size analysis due to missing data. We believe this in unlikely to cause substantial bias as the proportion is small and we do not believe missingness is related to the outcomes of interest. However, we cannot exclude the possibility.

Here, we have utilised a large brain tumour database linked to routine healthcare data to investigate the potential of earlier or delayed diagnosis to impact six important patient outcomes. Provided that we accept the logic that relates tumour size to earlier or later diagnosis, this study finds consistent evidence that delay does matter for brain tumour patients. New interventions leading to earlier diagnosis can hope to improve patient outcomes. Further studies, including prospective trials or real-world piloting, may be needed to confirm the impact of any specific early diagnosis intervention.

## Data Availability

Under data governance rules these patient data can be accessed by researchers via application to Public Health Scotland (PHS) electronic Data Research and Innovation Service (eDRIS).

## Author contributions

EG, MV, PH, and PB contributed to the conceptualization and study design. EG also contributed by writing of the original draft. PK and GT contributed to the data analysis. JC contributed to literature review. MB contributed though acquisition of funding. All authors contributed to data interpretation and writing through review and editing.

## Declaration of interests

EG is a paid consultant of Dxcover Ltd. JC, AL and MB are employees of Dxcover Ltd and own stock in Dxcover Ltd. Dxcover Ltd. LP is in receipt of honoraria from Dxcover Ltd. PB, PH, PK, GT and MH were supported in their work on this study from a contract between Dxcover Ltd and their employer (Edinburgh Innovations, University of Edinburgh).

## Source of Funding

Dxcover Ltd provided funding for data access, analysis and writing.

## Acknowledgements

We would like to thank patients for providing access to their data, allowing this study to be completed.

## Notes

### Author Declarations

As this is an observational study falling within the classification of service evaluation that uses secondary data previously collected in the course of routine patient care and therefore does not require patient consent provided that approval is granted the NHS Lothian DataLoch governance process that operates within NHS Lothian with authority delegated from the NHS Lothian Caldicott Guardian, NHS Research Ethics Service and the NHS Lothian Research and Development department. Data were accessed through eDRIS. The data were de-identified prior to use in the study.

