## Supplementary Appendix for "Does earlier diagnosis and treatment of brain tumours matter? Time-to-treatment intervals and tumour size at detection; impact on survival, recurrence, inpatient length of stay and neurological deficit"

### 1. Further information about variables included in the analysis

Brain tumour specific mortality definition: Cause of death matches any of these ICD-10 diagnoses: C700, C701, C709, C710, C711, C712, C713, C714, C715, C716, C717, C718, C719, C720, C721, C722,C723, C724, C725, C728, C729, C752, C753, C793, D320, D321, D329, D330, D331, D332, D333, D334, D339, D352, D354, D420, D429, D430, D431, D432, D433, D434, D444, D445.

### 2. Details of calculation of earlier detection example

A reduction in tumour size ($S_r$) due to earlier detection can be calculated from the size at detection ($S_d$) and an assumed exponential growth rate ($r$):

$$S_e = S_d \times \left(\frac{1}{1 + r^t}\right)$$

$$S_r = S_e - S_d$$

$S_e$ is the size at earlier detection and $t$ is time in days.

The average growth rate for GBM tumours has been estimated to be 1.2% (Swanton et al) and 2.2% (Feucht et al) per day, based on reported tumour volume or size doubling times. These two growth rate estimates were used in the example calculation.

The impact of an $S_r$ magnitude reduction in size on outcomes was calculated by applying the estimated OR/HR from regression analysis. reduction. For example, for outcomes such as cancer specific mortality with an estimated hazard ratio:

$$HR_{Sr} = e^{\log(HR)S_r}$$

In the example provided, a 60mm tumour with an average growth detected one month earlier would be expected to be between 17.5mm and 29.1mm smaller at that time. These reductions in size translate to hazard ratios of 0.82 and 0.72, equivalent to relative risk reductions of 18% and 28% respectively.

Relative risk reductions were translated to survival at 1 year and 3 years by assuming an exponential distribution of survival times, and survival for a tumour size of mean size (40mm) being approximately equal to mean survival.

The linear regression coefficients estimated for inpatient bed days were applied directly to the size reductions above. Cost savings were calculated using an inpatient bed day cost for a neurosurgical unit of £500 (Scottish Costs book).

### 3. Tumour size associations with patient outcomes (unadjusted)

Note the number of observations included in each analysis varies depending on the necessary variables and levels of missingness. The number of observation for each are reported in SA Table 21 to SA Table 23.

***SA Table 1: Association between tumour size and outcomes of interest (unadjusted) - overall***

| | **Outcome** | | | | | |
|---|---|---|---|---|---|---|
| | **30-day mortality** (n = 1070) | | **New or worsened neurological deficit** (n = 1070) | | **Inpatient bed days up to 12 months following diagnosis** (n = 949) | |
| **Predictor** | *Unadjusted OR (95% CI)* | *p-value* | *Unadjusted OR (95% CI)* | *p-value* | *Unadjusted Coefficient (SE)* | *p-value* |
| Tumour size (cm) | 1.21 (0.99 – 1.47) | 0.063 | 1.16 (1.07 – 1.27) | **<0.001*** | 1.50 (0.64) | **0.020*** |
| **Cont.** | | | | | | |
| | **Overall survival** (n = 1070) | | **Brain cancer specific survival** (n = 1070) | | **Recurrence** (n = 1070) | |
| **Predictor** | *Unadjusted HR (95% CI)* | *p-value* | *Unadjusted HR (95% CI)* | *p-value* | *Unadjusted HR (95% CI)* | *p-value* |
| Tumour size (cm) | 1.12 (1.08 – 1.17) | **<0.001*** | 1.19 (1.15 – 1.24) | **<0.001*** | 1.06 (1.00 – 1.12) | **0.037*** |

*(OR – Odds ratio, HR – Hazard ratio, CI – Confidence Interval, SE – Standard Error, * Statistically significant)*

***SA Table 2: Association between tumour size and outcomes of interest (unadjusted) – Glioblastoma subgroup***

| | **Outcome** | | | | | |
|---|---|---|---|---|---|---|
| | **30-day mortality** (n = 426) | | **New or worsened neurological deficit** (n = 426) | | **Inpatient bed days up to 12 months following diagnosis** (n = 398) | |
| **Predictor** | *Unadjusted OR (95% CI)* | *p-value* | *Unadjusted OR (95% CI)* | *p-value* | *Unadjusted Coefficient (SE)* | *p-value* |
| Tumour size (cm) | 1.49 (1.09 – 2.06) | **0.012*** | 0.98 (0.86 – 1.13) | 0.814 | 1.77 (1.05) | 0.093 |
| **Cont.** | | | | | | |
| | **Overall survival** (n = 426) | | **Brain cancer specific survival** (n = 426) | | **Recurrence** (n = 426) | |
| **Predictor** | *Unadjusted HR (95% CI)* | *p-value* | *Unadjusted HR (95% CI)* | *p-value* | *Unadjusted HR (95% CI)* | *p-value* |
| Tumour size (cm) | 1.11 (1.05 – 1.18) | **<0.001*** | 1.11 (1.05 – 1.18) | **<0.001*** | 0.89 (0.81 – 0.97) | **0.007*** |

*(OR – Odds ratio, HR – Hazard ratio, CI – Confidence Interval, SE – Standard Error, * Statistically significant)*

***SA Table 3: Association between tumour size and outcomes of interest (unadjusted) - Other glioma, other primary brain cancers, meningioma (Less aggressive primary brain tumours) subgroup***

| | Outcome | | | | | |
|---|---|---|---|---|---|---|
| | **30-day mortality** (n = 500) | | **New or worsened neurological deficit** (n = 500) | | **Inpatient bed days up to 12 months following diagnosis** (n = 418) | |
| **Predictor** | *Unadjusted OR (95% CI)* | *p-value* | *Unadjusted OR (95% CI)* | *p-value* | *Unadjusted Coefficient (SE)* | *p-value* |
| Tumour size (cm) | 0.95 (0.61 – 1.39) | 0.783 | 1.25 (1.10 – 1.42) | **0.001*** | 1.65 (0.97) | 0.091 |
| **Cont.** | | | | | | |
| | **Overall survival** (n = 500) | | **Brain cancer specific survival** (n = 500) | | **Recurrence** (n = 500) | |
| **Predictor** | *Unadjusted HR (95% CI)* | *p-value* | *Unadjusted HR (95% CI)* | *p-value* | *Unadjusted HR (95% CI)* | *p-value* |
| Tumour size (cm) | 1.09 (1.02 – 1.17) | **0.015*** | 1.12 (1.04 – 1.22) | **0.004*** | 1.16 (1.07 – 1.27) | **0.001*** |

*(OR – Odds ratio, HR – Hazard ratio, CI – Confidence Interval, SE – Standard Error, * Statistically significant)*

***SA Table 4: Association between tumour size and outcomes of interest (unadjusted) – Metastatic subgroup***

| | Outcome | | | | | |
|---|---|---|---|---|---|---|
| | **30-day mortality** (n = 144) | | **New or worsened neurological deficit** (n = 144) | | **Inpatient bed days up to 12 months following diagnosis** (n = 133) | |
| **Predictor** | *Unadjusted OR (95% CI)* | *p-value* | *Unadjusted OR (95% CI)* | *p-value* | *Unadjusted Coefficient (SE)* | *p-value* |
| Tumour size (cm) | 1.52 (0.89 – 2.58) | 0.115 | 0.97 (0.64 – 1.43) | 0.867 | 2.15 (2.32) | 0.357 |
| **Cont.** | | | | | | |
| | **Overall survival** (n = 144) | | **Brain cancer specific survival** (n = 144) | | **Recurrence** (n = 144) | |
| **Predictor** | *Unadjusted HR (95% CI)* | *p-value* | *Unadjusted HR (95% CI)* | *p-value* | *Unadjusted HR (95% CI)* | *p-value* |
| Tumour size (cm) | 1.22 (1.04 – 1.42) | **0.012*** | 1.14 (0.95 – 1.36) | 0.160 | 1.03 (0.82 – 1.30) | 0.771 |

*(OR – Odds ratio, HR – Hazard ratio, CI – Confidence Interval, SE – Standard Error, * Statistically significant)*

### 4. Time-to-treatment associations with patient outcomes (continuous specification, unadjusted and adjusted)

#### *SA Table 5: Association between the time-to-treatment window and outcomes of interest (unadjusted), Pooled*

| | Outcome | | | | | |
|---|---|---|---|---|---|---|
| | Overall survival | | Brain cancer-specific survival | | Recurrence | |
| **Predictor** | *Unadjusted HR (95% CI)* | *p-value* | *Unadjusted HR (95% CI)* | *p-value* | *Unadjusted HR (95% CI)* | *p-value* |
| Time radiological diagnosis to first treatment (days) | 0.9963 (0.9952 – 0.9974) | **<0.001*** | 0.9967 (0.9956 – 0.9979) | **<0.001*** | 0.9992 (0.9987 – 0.9998) | **0.006*** |
| Time from first presentation to treatment (days) | 0.9976 (0.9970 – 0.9982) | **<0.001*** | 0.9979 (0.9973 – 0.9985) | **<0.001*** | 0.9993 (0.9989 – 0.9997) | **0.001*** |
| **Cont.** | | | | | | |
| | New or worsened neurological deficit | | 30-day mortality | | | |
| **Predictor** | *Unadjusted OR (95% CI)* | *p-value* | *Unadjusted OR (95% CI)* | *p-value* | | |
| Time radiological diagnosis to first treatment (days) | 1.000 (1.000 – 1.001) | 0.718 | 0.930 (0.745 – 0.980) | 0.380 | | |
| Time from first presentation to treatment (days) | 1.000 (0.999 – 1.000) | 0.613 | 0.884 (0.728 – 0.990) | 0.110 | | |

#### *SA Table 6: Association between the time-to-treatment window and outcomes of interest (adjusted), Pooled*

| | Outcome | | | | | |
|---|---|---|---|---|---|---|
| | Overall survival | | Brain cancer-specific survival | | Recurrence | |
| **Predictor** | *Adjusted HR (95% CI)* | *p-value* | *Adjusted HR (95% CI)* | *p-value* | *Adjusted HR (95% CI)* | *p-value* |
| Time radiological diagnosis to first treatment (days) | 0.9979 (0.9971 – 0.9988) | **<0.001*** | 0.9987 (0.9979 – 0.9995) | **0.002*** | 0.9995 (0.9990 – 1.0000) | **0.050*** |
| Time from first presentation to treatment (days) | 0.9985 (0.9980 – 0.9991) | **<0.001*** | 0.9991 (0.9985 – 0.9996) | **0.001*** | 0.9995 (0.9991 – 0.9999) | **0.014*** |
| **Cont.** | | | | | | |

| | New or worsened neurological deficit | | 30-day mortality | |
|---|---|---|---|---|
| **Predictor** | *Adjusted OR (95% CI)* | *p-value* | *Adjusted OR (95% CI)* | *p-value* |
| Time radiological diagnosis to first treatment (days) | 1.000 (1.000 – 1.001) | 0.221 | 0.933 (0.741 – 0.986) | 0.431 |
| Time from first presentation to treatment (days) | 1.000 (1.000 – 1.001) | 0.627 | 0.881 (0.689 – 0.975) | 0.150 |

*(OR – Odds ratio, HR – Hazard ratio, CI – Confidence Interval, * Statistically significant)*

#### *SA Table 7: Association between the time-to-treatment window and outcomes of interest (unadjusted), Glioblastoma*

| | Outcome | | | | | |
|---|---|---|---|---|---|---|
| | Overall survival | | Brain cancer-specific survival | | Recurrence | |
| **Predictor** | *Unadjusted HR (95% CI)* | *p-value* | *Unadjusted HR (95% CI)* | *p-value* | *Unadjusted HR (95% CI)* | *p-value* |
| Time radiological diagnosis to first treatment (days) | 1.0011 (0.9987 – 1.0036) | 0.364 | 1.0013 (0.9988 – 1.0037) | 0.307 | 0.9962 (0.9903 – 1.0021) | 0.209 |
| Time from first presentation to treatment (days) | 1.0013 (0.9998 – 1.0028) | 0.081 | 1.0002 (0.9987 – 1.0017) | 0.808 | 0.9983 (0.9962 – 1.0005) | 0.129 |
| **Cont.** | | | | | | |
| | New or worsened neurological deficit | | 30-day mortality | | | |
| **Predictor** | *Unadjusted OR (95% CI)* | *p-value* | *Unadjusted OR (95% CI)* | *p-value* | | |
| Time radiological diagnosis to first treatment (days) | 0.995 (0.983 – 1.002) | 0.289 | 0.924 (0.632 – 1.009) | 0.555 | | |
| Time from first presentation to treatment (days) | 0.998 (0.994 – 1.001) | 0.193 | 0.855 (0.588 – 0.989) | 0.244 | | |

#### *SA Table 8: Association between the time-to-treatment window and outcomes of interest (adjusted), Glioblastoma*

| | Outcome | | | | | |
|---|---|---|---|---|---|---|
| | Overall survival | | Brain cancer-specific survival | | Recurrence | |
| **Predictor** | *Adjusted HR (95% CI)* | *p-value* | *Adjusted HR (95% CI)* | *p-value* | *Adjusted HR (95% CI)* | *p-value* |

| | | | | | | |
|---|---|---|---|---|---|---|
| Time radiological diagnosis to first treatment (days) | 1.0002 (0.9974 – 1.0030) | 0.886 | 1.0005 (0.9977 – 1.0032) | 0.736 | 0.9971 (0.9922 – 1.0021) | 0.260 |
| Time from first presentation to treatment (days) | 1.0012 (0.9998 – 1.0027) | 0.099 | 1.0001 (0.9985 – 1.0016) | 0.940 | 0.9986 (0.9965 – 1.0007) | 0.201 |
| **Cont.** | | | | | | |

| | New or worsened neurological deficit | | 30-day mortality | |
|---|---|---|---|---|
| **Predictor** | *Adjusted OR (95% CI)* | *p-value* | *Adjusted OR (95% CI)* | *p-value* |
| Time radiological diagnosis to first treatment (days) | 0.995 (0.983 – 1.001) | 0.275 | 0.908 (0.607 – 1.090) | 0.495 |
| Time from first presentation to treatment (days) | 0.998 (0.994 – 1.001) | 0.180 | 0.000 (0.000 – 5.043 x $10^{34}$) | 0.984 |

*(OR – Odds ratio, HR – Hazard ratio, CI – Confidence Interval, * Statistically significant)*

#### *SA Table 9: Association between the time-to-treatment window and outcomes of interest (unadjusted), Non-Glioblastoma Primary Brain Tumours*

| | Outcome | | | | | |
|---|---|---|---|---|---|---|
| | Overall survival | | Brain cancer-specific survival | | Recurrence | |
| **Predictor** | *Unadjusted HR (95% CI)* | *p-value* | *Unadjusted HR (95% CI)* | *p-value* | *Unadjusted HR (95% CI)* | *p-value* |
| Time radiological diagnosis to first treatment (days) | 0.9979 (0.9969 – 0.9990) | **<0.001*** | 0.9986 (0.9977 – 0.9995) | **0.003*** | 0.9996 (0.9992 – 1.0001) | 0.123 |
| Time from first presentation to treatment (days) | 0.9985 (0.9978 – 0.9991) | **<0.001*** | 0.9990 (0.9984 – 0.9996 | **0.001*** | 0.9996 (0.9992 – 1.0000) | 0.074 |
| **Cont.** | | | | | | |

| | New or worsened neurological deficit | | 30-day mortality | |
|---|---|---|---|---|
| **Predictor** | *Unadjusted OR (95% CI)* | *p-value* | *Unadjusted OR (95% CI)* | *p-value* |
| Time radiological diagnosis to first treatment (days) | 1.000 (1.000 – 1.001) | 0.178 | 1.000 (0.000 – 2.758 x $10^{39}$) | 1.000 |
| Time from first presentation to treatment (days) | 1.000 (1.000 – 1.001) | 0.568 | 1.000 (0.000 – 2.349 x $10^{33}$) | 1.000 |

#### *SA Table 10: Association between the time-to-treatment window and outcomes of interest (adjusted), Non-Glioblastoma Primary Brain Tumours*

| | **Outcome** | | | | | |
|---|---|---|---|---|---|---|
| | **Overall survival** | | **Brain cancer-specific survival** | | **Recurrence** | |
| **Predictor** | *Adjusted HR (95% CI)* | *p-value* | *Adjusted HR (95% CI)* | *p-value* | *Adjusted HR (95% CI)* | *p-value* |
| Time radiological diagnosis to first treatment (days) | 0.9979 (0.9969 – 0.9990) | **<0.001*** | 0.9987 (0.9978 – 0.9996) | **0.005*** | 0.9996 (0.9992 – 1.0001) | 0.117 |
| Time from first presentation to treatment (days) | 0.9985 (0.9979 – 0.9992) | **<0.001*** | 0.9991 (0.9985 – 0.9997) | **0.002*** | 0.9996 (0.9992 – 1.0000) | 0.053 |

**Cont.**

| | **New or worsened neurological deficit** | | **30-day mortality** | |
|---|---|---|---|---|
| **Predictor** | *Adjusted OR (95% CI)* | *p-value* | *Adjusted OR (95% CI)* | *p-value* |
| Time radiological diagnosis to first treatment (days) | 1.000 (1.000 – 1.001) | 0.187 | 1.000 (0.000 – 4.550 x $10^{39}$) | 1.000 |
| Time from first presentation to treatment (days) | 1.000 (1.000 – 1.001) | 0.557 | 1.000 (0.000 – 6.489 x $10^{33}$) | 1.000 |

*(OR – Odds ratio, HR – Hazard ratio, CI – Confidence Interval, * Statistically significant)*

#### *SA Table 11: Association between the time-to-treatment window and outcomes of interest (unadjusted), Metastatic tumours*

| | **Outcome** | | | | | |
|---|---|---|---|---|---|---|
| | **Overall survival** | | **Brain cancer-specific survival** | | **Recurrence** | |
| **Predictor** | *Unadjusted HR (95% CI)* | *p-value* | *Unadjusted HR (95% CI)* | *p-value* | *Unadjusted HR (95% CI)* | *p-value* |
| Time radiological diagnosis to first treatment (days) | 0.9995 (0.9979 – 1.0010) | 0.474 | 0.9976 (0.9917 – 1.0034) | 0.411 | 0.9965 (0.9882 – 1.0048) | 0.404 |
| Time from first presentation to treatment (days) | 0.9989 (0.9972 – 1.0007) | 0.249 | 0.9960 (0.9904 – 1.0016) | 0.163 | 0.9979 (0.9936 – 1.0021) | 0.324 |

**Cont.**

| | **New or worsened neurological deficit** | | **30-day mortality** | |
|---|---|---|---|---|
| **Predictor** | *Unadjusted OR (95% CI)* | *p-value* | *Unadjusted OR (95% CI)* | *p-value* |
| Time radiological | 0.999 | 0.698 | 0.924 (0.672 – 0.996) | 0.488 |

| | | | | |
|---|---|---|---|---|
| diagnosis to first treatment (days) | (0.984 – 1.003) | | | |
| Time from first presentation to treatment (days) | 1.000 (0.993 – 1.003) | 0.895 | 0.820 (0.485 – 0.977) | 0.224 |

*SA Table 12: Association between the time-to-treatment window and outcomes of interest (adjusted), Metastatic tumours*

| | Outcome | | | | | |
|---|---|---|---|---|---|---|
| | Overall survival | | Brain cancer-specific survival | | Recurrence | |
| **Predictor** | *Adjusted HR (95% CI)* | *p-value* | *Adjusted HR (95% CI)* | *p-value* | *Adjusted HR (95% CI)* | *p-value* |
| Time radiological diagnosis to first treatment (days) | 0.9998 (0.9982 – 1.0013) | 0.775 | 0.9995 (0.9947 – 1.0043) | 0.831 | 0.9955 (0.9855 – 1.0056) | 0.383 |
| Time from first presentation to treatment (days) | 0.9993 (0.9975 – 1.0012) | 0.475 | 0.9974 (0.9921 – 1.0027) | 0.330 | 0.9978 (0.9933 – 1.0023) | 0.335 |
| **Cont.** | | | | | | |
| | **New or worsened neurological deficit** | | **30-day mortality** | | | |
| **Predictor** | *Adjusted OR (95% CI)* | *p-value* | *Adjusted OR (95% CI)* | *p-value* | | |
| Time radiological diagnosis to first treatment (days) | 0.998 (0.980 – 1.003) | 0.632 | 1.015 (0.708 – 1.409) | 0.917 | | |
| Time from first presentation to treatment (days) | 0.999 (0.992 – 1.003) | 0.770 | 0.877 (0.475 – 1.039) | 0.488 | | |

*(OR – Odds ratio, HR – Hazard ratio, CI – Confidence Interval, * Statistically significant)*

### 5. Time-to-treatment associations with patient outcomes, stratified by time-to-treatment category (continuous specification, unadjusted and adjusted)

*SA Table 13: Association between the time-to-treatment window (weeks) and outcomes of interest (unadjusted) – Time-to-treatment ≤ 2 weeks*

| | Outcome | | | | | |
|---|---|---|---|---|---|---|
| | Overall survival | | Brain cancer-specific survival | | Recurrence | |
| **Predictor** | *Unadjusted HR (95% CI)* | *p-value* | *Unadjusted HR (95% CI)* | *p-value* | *Unadjusted HR (95% CI)* | *p-value* |

| | | | | | | |
|---|---|---|---|---|---|---|
| Time radiological diagnosis to first treatment (weeks) | 1.1472 (0.9186 – 1.4327) | 0.226 | 1.1411 (0.9010 – 1.4451) | 0.273 | 0.8877 (0.6839 – 1.1522) | 0.371 |
| Time from first presentation to treatment (weeks) | 1.1133 (0.6504 – 1.9058) | 0.695 | 1.2108 (0.6784 – 2.1610) | 0.518 | 0.7607 (0.4040 – 1.4324) | 0.397 |
| **Cont.** | | | | | | |

| | New or worsened neurological deficit | | 30-day mortality | |
|---|---|---|---|---|
| **Predictor** | *Unadjusted OR (95% CI)* | *p-value* | *Unadjusted OR (95% CI)* | *p-value* |
| Time radiological diagnosis to first treatment (weeks) | 1.6205 (1.0487 – 2.5406) | **0.032*** | 4.9652 (0.3163 – 564.2627) | 0.348 |
| Time from first presentation to treatment (weeks) | 1.4306 (0.4559 – 5.1605) | 0.556 | 11.9550 (0.1026 – 553972.4311) | 0.479 |

#### *SA Table 14: Association between the time-to-treatment window (weeks) and outcomes of interest (adjusted) – Time-to-treatment ≤ 2 weeks*

| | Outcome | | | | | |
|---|---|---|---|---|---|---|
| | Overall survival | | Brain cancer-specific survival | | Recurrence | |
| **Predictor** | *Adjusted HR (95% CI)* | *p-value* | *Adjusted HR (95% CI)* | *p-value* | *Adjusted HR (95% CI)* | *p-value* |
| Time radiological diagnosis to first treatment (weeks) | 1.0148 (0.8032 – 1.2821) | 0.902 | 0.9727 (0.7571 – 1.2498) | 0.829 | 0.8152 (0.6249 – 1.0634) | 0.132 |
| Time from first presentation to treatment (weeks) | 0.9685 (0.5013 – 1.8712) | 0.924 | 1.1344 (0.5669 – 2.2702) | 0.722 | 0.6666 (0.3019 – 1.4720) | 0.316 |
| **Cont.** | | | | | | |

| | New or worsened neurological deficit | | 30-day mortality | |
|---|---|---|---|---|
| **Predictor** | *Adjusted OR (95% CI)* | *p-value* | *Adjusted OR (95% CI)* | *p-value* |
| Time radiological diagnosis to first treatment (weeks) | 1.5156 (0.9620 – 2.4199) | 0.076 | 5.7011 (0.3247 – 822.4359) | 0.325 |
| Time from first presentation to treatment (weeks) | 0.8835 (0.1892 – 4.2664) | 0.873 | $3.765 \times 10^{-23}$ (0.0000 – Inf) | 0.999 |

#### *SA Table 15: Association between the time-to-treatment window (weeks) and outcomes of interest (unadjusted) – Time-to-treatment 2-6 weeks*

| | Outcome | | | | | |
|---|---|---|---|---|---|---|
| | Overall survival | | Brain cancer-specific survival | | Recurrence | |
| **Predictor** | *Unadjusted HR (95% CI)* | *p-value* | *Unadjusted HR (95% CI)* | *p-value* | *Unadjusted HR (95% CI)* | *p-value* |
| Time radiological diagnosis to first treatment (weeks) | 0.9194 (0.8247 – 1.0251) | 0.130 | 0.8871 (0.7914 – 0.9943) | **0.040*** | 0.8580 (0.7399 – 0.9950) | **0.043*** |
| Time from first presentation to treatment (weeks) | 1.0110 (0.8974 – 1.1389) | 0.857 | 0.9857 (0.8666 – 1.1211) | 0.826 | 0.8803 (0.7570 – 1.0237) | 0.098 |
| **Cont.** | | | | | | |
| | New or worsened neurological deficit | | 30-day mortality | | | |
| **Predictor** | *Unadjusted OR (95% CI)* | *p-value* | *Unadjusted OR (95% CI)* | *p-value* | | |
| Time radiological diagnosis to first treatment (weeks) | 0.9642 (0.7738 – 1.1959) | 0.742 | 1.0000 (0.0000 – Inf) | 1.000 | | |
| Time from first presentation to treatment (weeks) | 0.9743 (0.7688 – 1.2345) | 0.829 | 0.0000 (NA – Inf) | 0.994 | | |

#### *SA Table 16: Association between the time-to-treatment window (weeks) and outcomes of interest (adjusted) – Time-to-treatment 2-6 weeks*

| | Outcome | | | | | |
|---|---|---|---|---|---|---|
| | Overall survival | | Brain cancer-specific survival | | Recurrence | |
| **Predictor** | *Adjusted HR (95% CI)* | *p-value* | *Adjusted HR (95% CI)* | *p-value* | *Adjusted HR (95% CI)* | *p-value* |
| Time radiological diagnosis to first treatment (weeks) | 0.9667 (0.8584 – 1.0887) | 0.576 | 0.9714 (0.8581 – 1.0997) | 0.647 | 0.8702 (0.7468 – 1.0139) | 0.075 |
| Time from first presentation to treatment (weeks) | 1.0820 (0.9596 – 1.2200) | 0.198 | 1.0410 (0.9171 – 1.1816) | 0.534 | 0.8915 (0.7632 – 1.0414) | 0.148 |
| **Cont.** | | | | | | |
| | New or worsened neurological deficit | | 30-day mortality | | | |
| **Predictor** | *Adjusted OR (95% CI)* | *p-value* | *Adjusted OR (95% CI)* | *p-value* | | |
| Time radiological diagnosis to first treatment (weeks) | 0.9957 (0.7930 – 1.2460) | 0.970 | 1.0000 (0.0000 – Inf) | 1.000 | | |

| | | | | |
|---|---|---|---|---|
| Time from first presentation to treatment (weeks) | 1.0084 (0.7893 – 1.2891) | 0.947 | 0.0000 (0.0000 – Inf) | 0.994 |

#### *SA Table 17: Association between the time-to-treatment window (weeks) and outcomes of interest (unadjusted) – Time-to-treatment 6-12 weeks*

| | **Outcome** | | | | | |
|---|---|---|---|---|---|---|
| | **Overall survival** | | **Brain cancer-specific survival** | | **Recurrence** | |
| **Predictor** | *Unadjusted HR (95% CI)* | *p-value* | *Unadjusted HR (95% CI)* | *p-value* | *Unadjusted HR (95% CI)* | *p-value* |
| Time radiological diagnosis to first treatment (weeks) | 0.7479 (0.6224 – 0.8987) | **0.002*** | 0.7613 (0.6199 – 0.9351) | **0.009*** | 0.9941 (0.8296 – 1.1912) | 0.949 |
| Time from first presentation to treatment (weeks) | 0.8993 (0.8138 – 0.9939) | **0.037*** | 0.9360 (0.8424 – 1.0401) | 0.219 | 0.9059 (0.8012 – 1.0243) | 0.115 |
| **Cont.** | | | | | | |
| | **New or worsened neurological deficit** | | **30-day mortality** | | | |
| **Predictor** | *Unadjusted OR (95% CI)* | *p-value* | *Unadjusted OR (95% CI)* | *p-value* | | |
| Time radiological diagnosis to first treatment (weeks) | 1.0164 (0.7559 – 1.3489) | 0.911 | 1.0000 (0.0000 – Inf) | 1.000 | | |
| Time from first presentation to treatment (weeks) | 0.8661 (0.7044 – 1.0560) | 0.162 | 1.0000 (0.0000 – Inf) | 1.000 | | |

#### *SA Table 18: Association between the time-to-treatment window (weeks) and outcomes of interest (adjusted) – Time-to-treatment 6-12 weeks*

| | **Outcome** | | | | | |
|---|---|---|---|---|---|---|
| | **Overall survival** | | **Brain cancer-specific survival** | | **Recurrence** | |
| **Predictor** | *Adjusted HR (95% CI)* | *p-value* | *Adjusted HR (95% CI)* | *p-value* | *Adjusted HR (95% CI)* | *p-value* |
| Time radiological diagnosis to first treatment (weeks) | 0.8075 (0.6537 – 0.9974) | **0.047*** | 0.7876 (0.6236 – 0.9947) | **0.045*** | 1.0733 (0.8730 – 1.3196) | 0.502 |
| Time from first presentation to treatment (weeks) | 0.9561 (0.8596 – 1.0634) | 0.408 | 0.9908 (0.8858 – 1.1084) | 0.872 | 0.9150 (0.8057 – 1.0390) | 0.171 |
| **Cont.** | | | | | | |
| | **New or worsened neurological deficit** | | **30-day mortality** | | | |

| Predictor | *Adjusted OR (95% CI)* | *p-value* | *Adjusted OR (95% CI)* | *p-value* |
|---|---|---|---|---|
| Time radiological diagnosis to first treatment (weeks) | 1.0479 (0.7512 – 1.4548) | 0.779 | 1.0000 (0.0000 – Inf) | 1.000 |
| Time from first presentation to treatment (weeks) | 0.8618 (0.6912 – 1.0640) | 0.174 | 1.0000 (0.0000 – Inf) | 1.000 |

#### *SA Table 19: Association between the time-to-treatment window (weeks) and outcomes of interest (unadjusted) – Time-to-treatment > 12 weeks*

| | **Outcome** | | | | | |
|---|---|---|---|---|---|---|
| | **Overall survival** | | **Brain cancer-specific survival** | | **Recurrence** | |
| **Predictor** | *Unadjusted HR (95% CI)* | *p-value* | *Unadjusted HR (95% CI)* | *p-value* | *Unadjusted HR (95% CI)* | *p-value* |
| Time radiological diagnosis to first treatment (weeks) | 0.9942 (0.9889 – 0.9996) | **0.035*** | 0.9946 (0.9889 – 1.0003) | 0.062 | 0.9962 (0.9921 – 1.0003) | 0.066 |
| Time from first presentation to treatment (weeks) | 0.9895 (0.9850 – 0.9940) | **<0.001*** | 0.9912 (0.9867 – 0.9957) | **<0.001*** | 0.9966 (0.9935 – 0.9997) | **0.030*** |

**Cont.**

| | **New or worsened neurological deficit** | | **30-day mortality** | |
|---|---|---|---|---|
| **Predictor** | *Unadjusted OR (95% CI)* | *p-value* | *Unadjusted OR (95% CI)* | *p-value* |
| Time radiological diagnosis to first treatment (weeks) | 1.0039 (0.9995 – 1.0084) | 0.075 | 1.0000 (0.0000 – Inf) | 1.000 |
| Time from first presentation to treatment (weeks) | 1.0007 (0.9967 – 1.0042) | 0.717 | 1.0000 (0.0000 – ~Inf) | 1.000 |

#### *SA Table 20: Association between the time-to-treatment window (weeks) and outcomes of interest (adjusted) – Time-to-treatment > 12 weeks*

| | **Outcome** | | | | | |
|---|---|---|---|---|---|---|
| | **Overall survival** | | **Brain cancer-specific survival** | | **Recurrence** | |
| **Predictor** | *Adjusted HR (95% CI)* | *p-value* | *Adjusted HR (95% CI)* | *p-value* | *Adjusted HR (95% CI)* | *p-value* |
| Time radiological diagnosis to first treatment (weeks) | 0.9940 (0.9884 – 0.9996) | **0.035*** | 0.9950 (0.9893 – 1.0006) | 0.081 | 0.9961 (0.9921 – 1.0001) | 0.058 |

| | | | | | | |
|---|---|---|---|---|---|---|
| Time from first presentation to treatment (weeks) | 0.9937<br>(0.9896 – 0.9978) | **0.003*** | 0.9957<br>(0.9917 – 0.9998) | **0.038*** | 0.9969<br>(0.9939 – 1.0000) | 0.049 |

**Cont.**

| | **New or worsened neurological deficit** | | **30-day mortality** | |
|---|---|---|---|---|
| **Predictor** | *Adjusted OR (95% CI)* | *p-value* | *Adjusted OR (95% CI)* | *p-value* |
| Time radiological diagnosis to first treatment (weeks) | 1.0043<br>(0.9997 – 1.0089) | 0.058 | 1.0000<br>(0.0000 – Inf) | 1.000 |
| Time from first presentation to treatment (weeks) | 1.0014<br>(0.9973 – 1.0051) | 0.475 | 1.0000<br>(0.0000 – ~Inf) | 1.000 |

### 6. Number of Observations in Regression Analysis

Note the number of observations included in each analysis varies depending on the necessary variables and levels of missingness.

***SA Table 21: Number of Observations, Tumour size associations with patient outcomes***

| | **All patients** | | **GBM** | | **Non-GBM** | | **Metastasis** | |
|---|---|---|---|---|---|---|---|---|
| | Unadj. | Adj. | Unadj. | Adj. | Unadj. | Adj. | Unadj. | Adj. |
| All-cause mortality<br>Brain cancer mortality<br>Recurrence<br>30-day mortality<br>Neuro deficit | 1070 | 1064 | 426 | 425 | 500 | 496 | 144 | 143 |
| Inpatient bed days (12 months following diag.) | 949 | 944 | 398 | 397 | 418 | 415 | 133 | 132 |

***SA Table 22: Number of Observations, Time-to-treatment associations with patient outcomes (Diagnosis to treatment***

| | **All patients** | | **GBM** | | **Non-GBM** | | **Metastasis** | |
|---|---|---|---|---|---|---|---|---|
| | Unadj. | Adj. | Unadj. | Adj. | Unadj. | Adj. | Unadj. | Adj. |
| All-cause mortality<br>Brain cancer mortality<br>Recurrence<br>30-day mortality | 911 | 905 | 355 | 354 | 453 | 449 | 103 | 102 |

| Neuro deficit |
|---|

*SA Table 23: Number of Observations, Time-to-treatment associations with patient outcomes - (First presentation to treatment)*

| **Time-to-treatment associations with patient outcomes - (First presentation to treatment**) | **All patients** | | **GBM** | | **Non-GBM** | | **Metastasis** | |
|---|---|---|---|---|---|---|---|---|
| | Unadj. | Adj. | Unadj. | Adj. | Unadj. | Adj. | Unadj. | Adj. |
| All-cause mortality<br>Brain cancer mortality<br>Recurrence<br>30-day mortality<br>Neuro deficit | 872 | 867 | 343 | 342 | 431 | 428 | 98 | 97 |

### 7.Supplemenary Figures

***Figure SA1 - Forest Plot with Subgroups, Main Outcomes***

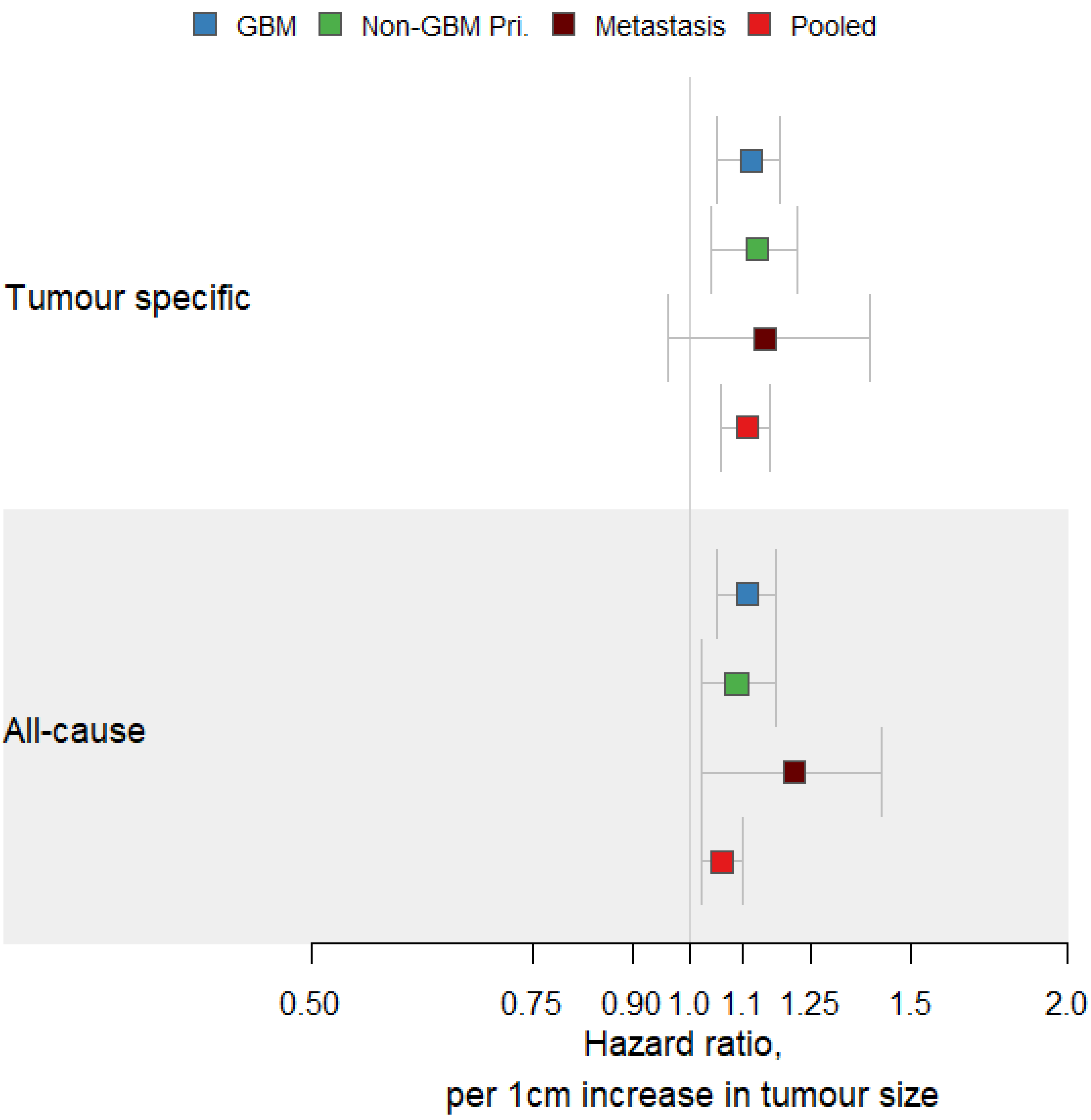
